# Association of COVID-19 RT-qPCR test false-negative rate with patient age, sex and time since diagnosis

**DOI:** 10.1101/2020.10.30.20222935

**Authors:** Matan Levine-Tiefenbrun, Idan Yelin, Hedva Uriel, Jacob Kuint, Licita Schreiber, Esma Herzel, Rachel Katz, Amir Ben-Tov, Tal Patalon, Gabriel Chodick, Roy Kishony

**Author notes:** These authors contributed equally to this work (Matan Levine-Tiefenbrun, Idan Yelin, Hedva Uriel).

## Abstract

**Background:** Routine testing for SARS-CoV-2 in the community is essential for guiding key epidemiological decisions from the quarantine of individual patients to enrolling regional and national preventive measures. Yet, the primary testing tool, the RT-qPCR based testing, is notoriously known for its low sensitivity, i.e. high risk of missed detection of carriers. Quantifying the false-negative rate (FNR) of the RT-qPCR test at the community settings and its dependence on patient demographic and disease progression is therefore key in designing and refining strategies for disease spread prevention.

**Methods:** Analyzing 843,917 test results of 521,696 patients, we identified false-negative (FN) and true-positive (TP) results as negative and positive results preceded by a COVID-19 diagnosis and followed by a later positive test. Regression analyses were used to determine associations of false-negative results with time of sampling after diagnosis, patient demographics and viral loads based on RT-qPCR Ct values of the next positive tests.

**Findings:** The overall FNR was 22.8%, which is consistent with previous studies. Yet, this rate was much lower at the first 5 days following diagnosis (10.7%) and only increased in later dates. Furthermore, the FNR was strongly associated with demographics, with odds ratio of 1.74 (95% CI: 1.58-1.90) for women over men and 1.36 (95% CI: 1.34-1.39) for 10 years younger patients. Finally, FNR was associated with viral loads (p-value 0.0005), with a difference of 1.50 (95% CI: 0.70-2.30) between the average Ct of the N gene in a positive test following a false-negative compared to a positive test following a true-positive.

**Interpretation:** Our results show that in the first few days following diagnosis, when results are critical for quarantine decisions, RT-qPCR testing is more reliable than previously reported. Yet the reliability of the test result is reduced in later days as well as for women and younger patients, where the viral loads are typically lower.

**Funding:** This research was supported by the ISRAEL SCIENCE FOUNDATION (grant No. 3633/19) within the KillCorona – Curbing Coronavirus Research Program.

## Introduction

The ongoing COVID-19 pandemic has already infected more than 45 million people worldwide (https://coronavirus.jhu.edu/map.html, October 30th, 2020^1^). A major tool in combating the pandemic is testing for viral carriage, which is used for both diagnostic and epidemiologic purposes. The most commonly used viral detection tests are based on the reverse transcription quantitative polymerase chain reaction of viral genes (RT-qPCR). This nucleic acid test is of high specificity, i.e. very low false-positive rate^2–5^. In contrast, a high false-negative rate was reported for these tests^6–10^. These high false-negative rates impede local and global efforts to slow down disease spread, as patients incorrectly diagnosed as non-carriers may subsequently infect additional people^11^. Systematically quantifying the rate of false-negative results and its dependencies on disease progression and patient demographics is critical for disease spread modeling, public health policy making and person-level quarantine decisions.

Various approaches have been taken to estimate the false-negative rate of COVID-19 RT-qPCR tests. Measuring the rate of false-negative results in a population of patients with highly specific pathologies (*e*.*g*. chest CT), has initially alerted physicians and epidemiologists of the high false-negative rate, estimated at approximately 30% ^4–7,9^. A meta-analysis of multiple such studies found that the reported rates were highly variable with a mean false-negative rate of 11%^12^. However, and as previously noted^12,13^, these meta-analysis studies were necessarily based on a combination of variable studies of non-uniform origins and methodologies, typically involving small groups of patients. A more recent systematic approach was based on ‘longitudinal testing’ in which the accuracy of each test is determined based on later tests of the same patient: a negative test which is directly followed by a positive one is deemed false negative. Application of this approach in a hospital setting resulted in an estimation of a false-negative rate of 17.8% ^13^. Systematic large scale studies at the community, critical for epidemiological disease control, have been lacking.

Beyond the average false negative rate, it is also important to understand whether and how the false-negative rate is associated with patient-specific and sample-specific attributes. Meta-analysis studies showed a strong association of false-negative results with time since exposure^14^ or time since onset of symptoms^15^. At the patient specific level, as viral load is associated with time since onset of symptoms, sex and age^16–22^, it has been proposed that false-negative rates might also depend on demographics, but current studies lacked statistical power for quantifying such dependencies^23^.

Here, we apply a longitudinal testing based approach to a large dataset of patient-level test series with linked demographics and electronic health records, to qunatify the false-negative rate of COVID-19 test results at the community and its associations with age, sex and time since diagnosis. Finally, we test whether the risk of false-negative results is associated with viral load at the single-patient level.

## Methods

### Data collection

Anonymized clinical records of SARS-CoV-2 RT-qPCR test results (test reports) were retrieved by Maccabi Healthcare Services (MHS) for the period between February 8th and September 24th 2020. Records of COVID-19 or COVID-19-related diagnoses by physicians (diagnosis reports) and referrals based on suspected exposure to the disease (epidemiological-based referrals) were retrieved for these patients. When available, fluorescence measurements data of the PCR test were retrieved for each test (*RT-qPCR measurements*). Randomly generated identifiers were used to link between test results and diagnosis codes.

#### Test results

MHS aggregates all test results for all its members, whether or not the test itself was performed by MHS laboratory. Test results data included, for each test: random patient number, sample number, sample execution date and test result. Test results were either “positive” (7.4%), “negative” (92%) or “borderline-positive” (0.6%, which we considered as positive in our analysis). Patients for whom two tests with different results were recorded on the same day were excluded from the analysis (274 patients, 0.05%).

#### Diagnosis reports

Diagnoses are routinely recorded in MHS database. For all patients with at least one positive test result, we retrieved any symptom-based COVID-19 diagnoses recorded prior to their first test. Diagnosis report data included: random patient number, diagnosis date, and diagnosis code (ICD9 and internal MHS codes).

#### Epidemiologically-based referrals

Since April 3rd 2020, an epidemiologically-based referral was filled by physicians referring a patient to a SARS-CoV-2 RT-qPCR test. Each referral report included: random patient number, referral number, referral date and referral cause.

#### RT-qPCR measurements

For each test, the following data were included: sample number, PCR machine number, test well number, test date, Ct values for 4 channels: FAM, Cal Red 610 Quasar 670 and HEX, corresponding to the measurements of E gene, RdRp gene, N gene and the internal control, respectively.

### Assigning patient diagnosis date

For each patient, the earliest date of COVID-19 symptoms and/or epidemiologically-based referral was considered as “date of diagnosis”. When both symptom-based diagnosis and epidemiological-based referrals were available, they were usually recorded on the same day. For simplicity, we excluded a small number of patients for whom both a diagnosis and a referral were available, but were more than a day apart (5.2% of diagnosed patients).

### Calculating the false negative rate

For any patient with at least one positive test, a ‘positive period’ was defined as the period between their date of diagnosis and their last positive test. Negative test reports during this period were regarded as false-negative (FN), while positive test reports during this period were regarded as true-positive (TP). False-negative rate (FNR) was calculated as 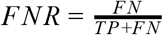.

### Logistic regression

Logistic regression of a false-negative versus true-positive result was performed using Python’s statsmodels library. The probability of a false-negative result was fitted to the test result (true-positive: 1, false-negative: 0) for all tests within the positive period.

### Linear regression

Linear regression of Ct values for each fluorescence channel was performed using Python’s statsmodels library.

### Calculating odds ratios from logistic regression

Odds ratios (OR) were calculated from the coefficients of the above logistic regression. For the binary variable sex (male: 1, female: 0), OR was defined as: *OR*_*sex*_ = *exp*(*C*_*sex*_), while for the continuous variables age and day, ORs were defined as: *exp*(*C*_*age*_ * *age*_*older*_− *C*_*age*_* *age*_*younger*_) younger versus older, *exp*(*C*_*day*_ * *day*_*early*_ − *C*_*day*_ * *day*_*late*_) early versus late, where *C*_*sex*_, *C*_*day*_, *C*_*age*_ are the coefficients for sex, day and age variables,respectivley.

### Curve fitting for FNR over time

Test results were grouped by day since diagnosis. FNR was calculated separately for each group. Data was fitted by 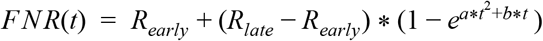, where *t* is time since diagnosis, and *a, b*, R_*early*_ and R_*late*_ are the fitted parameters, with the latter two standing for the false negative rates at the early and late phases, respectively.

### Differences in FNR between age groups

According to patient age, test results were divided into two groups of similar size (<40, ≥40 years). FNR was calculated separately for each group. Statistical significance for differences in FNR between groups was tested using a two-sided Fisher’s exact test (SciPy in Python).

### Ethical approval

The study protocol was approved by the ethics committee of Maccabi Healthcare Services, Tel-Aviv, Israel. IRB number: 0066-20-MHS.

## Results

Among all ∼2 million MHS patients, we identified 843,917 recorded tests for 521,696 patients (table 1). Within this set, 51,499 patients had at least one positive result. As quarantine discharge policy was based on test results, patients were often repeatedly tested, resulting in a series of test results for each patient. In our analysis, we focused on 7,872 patients with well-defined test series, satisfying the following conditions: (1) had a defined diagnosis date; (2) had at least one positive sample within 14 days following the diagnostic date; (3) had a test series that ended with a negative result (table 1). The vast majority of these test series ended with 2 or more negative results, in agreement with the discharge policy (68% of patients, supplementary table 1).

**Table 1.** Study population characteristics^*^.

| Characteristic | Study population<br>(N=7,872) |
| --- | --- |
| Age |  |
| Mean - yr | 36.78±20.15 |
| Distribution - no. (%) |  |
| <40 yr | 4,401 (55.90) |
| ≥40 yr | 3,471 (44.10) |
| Sex - no. (%) |  |
| Male | 4,164 (52.90) |
| Female | 3,708 (47.10) |
| Test result (%) |  |
| Positive | 14,588 (45.23) |
| Negative | 17,668 (54.77) |
| Median positive period length - days (IQR) | 5 (1-16) |
| Median time to recovery - days (IQR)** | 18 (13-24) |
| Median test series length - days (IQR) | 22 (16-30) |

\*Plus-minus values are means ±SD. IQR denotes interquartile range
\*\*For patients ending with negative

False negative and true positive test results were defined based on their context within a patient series of test results. We considered, for each patient, the series of test results following diagnosis (figure 1A,B). The median day since diagnosis until the first negative result, indicating recovery, was 18 (IQR: 13-24; table 1). For some patients a negative result came only after more than 50 days (2.4% of patients). For each patient, we then consider the period from diagnosis date to the last positive sample as a period in which the patient is carrying the virus, even if not at detectable loads yet (“positive period”). Taking an epidemiological stand, negative test results within this patient-specific positive time period were regarded as false-negative (FN) results. Similarly, positive test results within this period were regarded as true-positive (TP) (figure 1A). To avoid bias for true-positive results, the last positive result, used for marking the end of the positive period, was not counted towards TP. The rate of negative results increased over time, and at 20 days after diagnosis the number of negative tests first surpassed the number of positive or false-negative results (figure 1B,C). In total, we identified 1,982 test results defined as FN and 6,715 test results defined as TP, indicating an overall false-negative rate of 22.8% which is within the wide range of previously reported FNR^13,23–28^.

**Figure 1.**
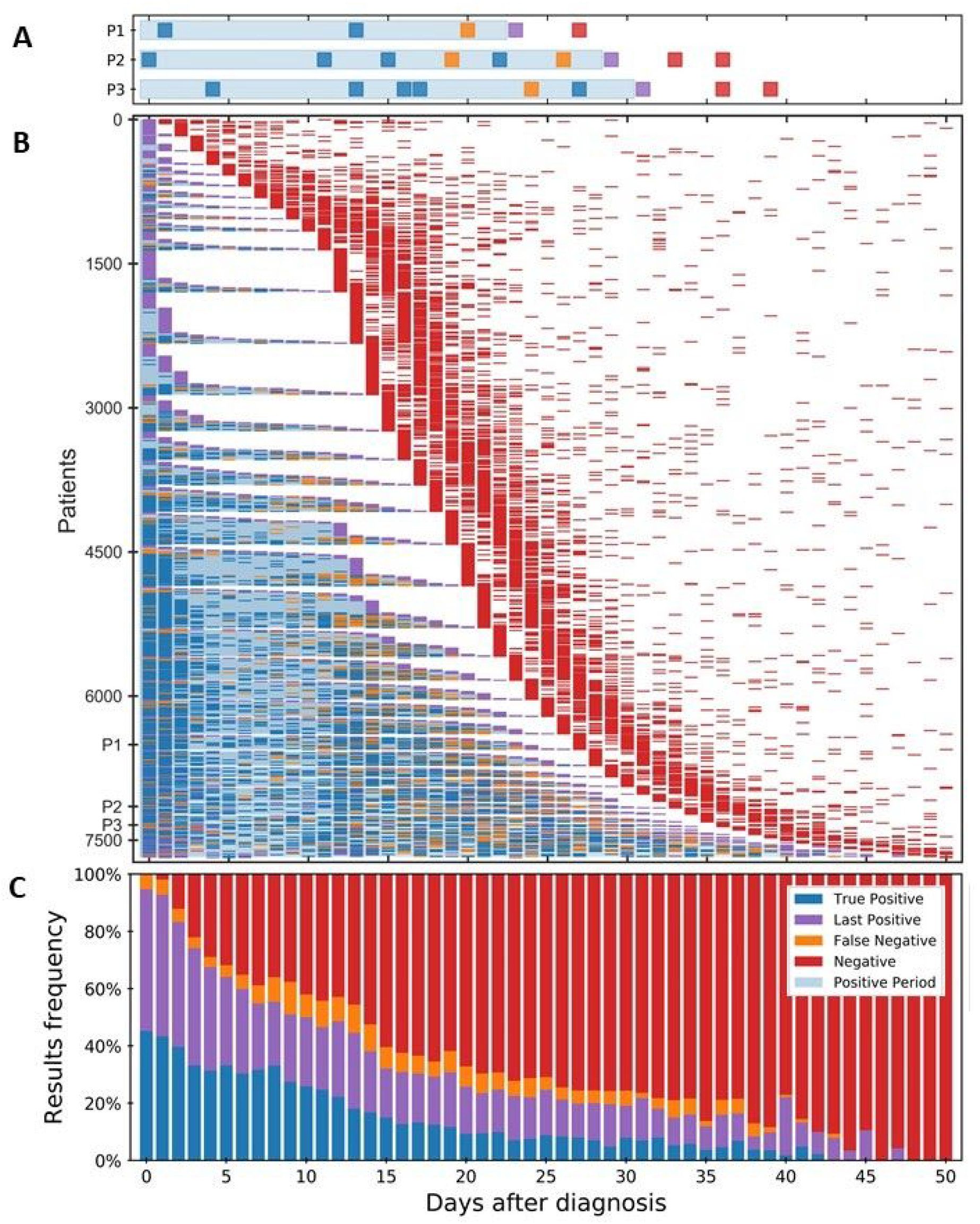
Longitudinal RT-qPCR SARS-CoV-2 test results for diagnosed COVID-19 patients. (**A**) Test results for 3 representative patients (P1, P2 and P3). The day of diagnosis and the last positive test result (purple) demarcate the ‘positive period’ (light blue shading). Negative tests within this individually determined period were regarded as ‘false negative’ (orange). Similarly, Positive tests within the ‘positive period’ were regarded as ‘true positive’ (blue). All test series end with a sequence of one or more negative results (red). (**B**) Longitudinal SARS-CoV-2 test results for the study population (table 1; for clarity, 191 patients for whom the first negative sample (red) was obtained more than 50 days after the day of diagnosis, were omitted). Patients are sorted by the dates, relative to diagnosis, of their first negative result, then by the relative date of the last positive result. (**C**) Frequency of test results per day relative to diagnosis.

To identify personalized features associated with false-negative results, we performed multivariate logistic regression for the odds of a false-negative versus true-positive result (Methods: *‘Logistic regression’* and *‘Calculating odds ratio from logistic regression’*). Patient age, sex and number of days from day of diagnosis were all associated with a false-negative result (supplementary table 2). Patient age was strongly anti-correlated with a false-negative result, with odds ratio of 1.36 (95% CI: 1.34-1.39) for 10 years younger patients. The number of days from the day of diagnosis was positively correlated with a false-negative result, with OR of 2.08 (95% CI: 1.85-2.33) for samples taken at day 15 compared to samples taken at day 0. Lastly, patient sex was also associated with false-negative results, with female to male odds ratio of 1.74 (95% CI: 1.58-1.9).

Following the observed association between time after diagnosis and false-negative result, we characterized the FNR during disease progression. Calculating FNR per day after diagnosis (Methods: *‘calculating FNR’*), we found that FNR followed 3 distinct phases: at the first few days following diagnosis, it was fairly constant and low (10.7%, days 0-5). It then gradually increased over days 6-15, and finally it plateaued at high rates of about 39% (Methods: *‘Curve fitting for FNR over time’*; figure 2A).

**Figure 2.**
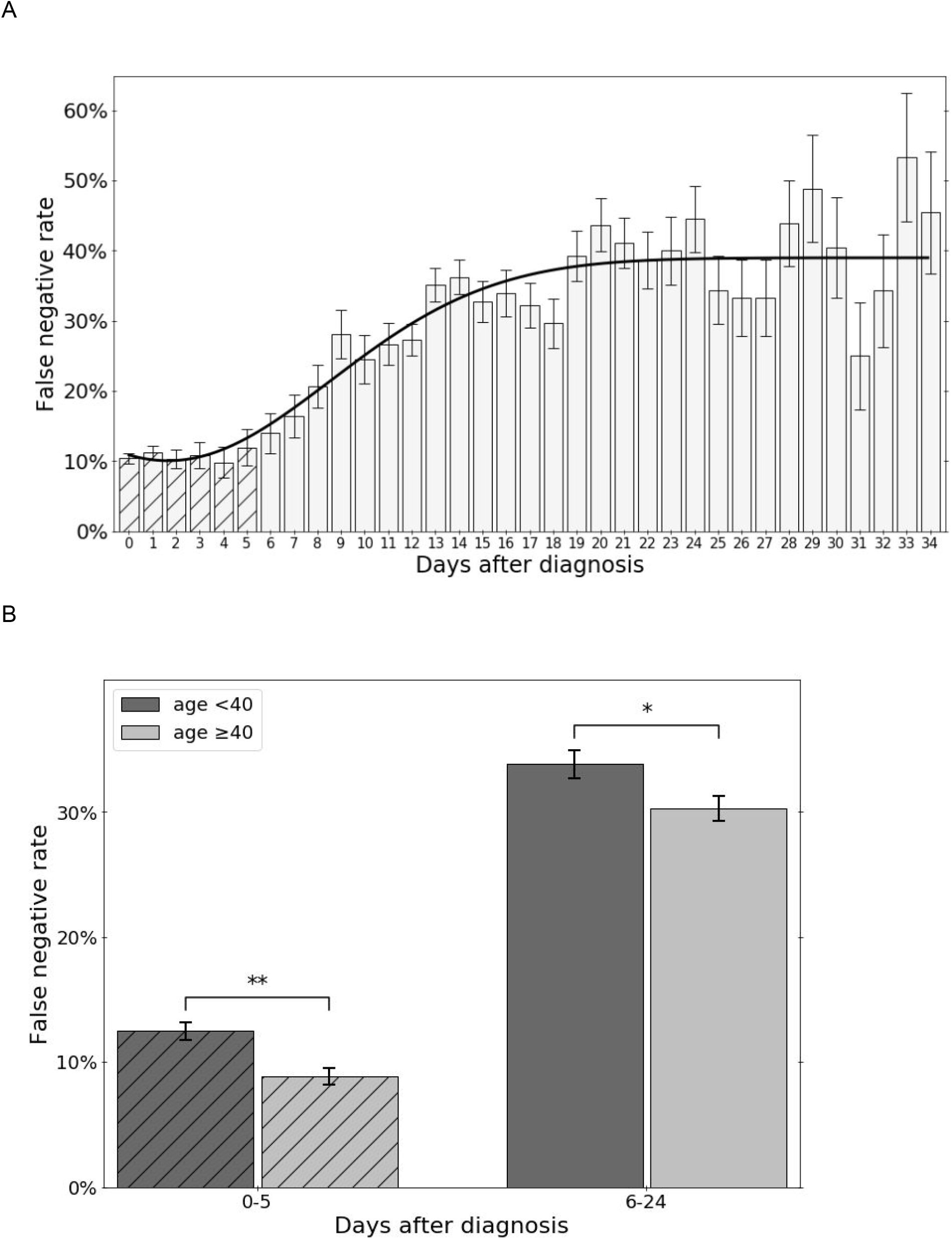
False-negative rate changes along time after day of diagnosis and differs between age groups. FNR per day after diagnosis was calculated for 8,697 tests. (**A**) daily FNR for days between day of diagnosis (day 0) to 34 days after diagnosis. FNR is fitted with 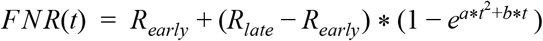. (**B**) Difference in FNR between two age groups (<40 and ≥40, dark and light grey, respectively) calculated separately for early and late days after diagnosis (hatched and empty, respectively). Fisher exact test (Methods: ‘Differences in FNR between age groups’). * - p-value<0.05, ** - p-value<0.01. Error bars indicate SD.

We next focused on the earlier days after diagnosis (days 0-5), in which FNR was relatively low, and in which a precise diagnosis is most critical for epidemiological patient-level quarantine control. Multivariate logistic regression analysis of test results for these days alone identified an association of false-negative results during these days with sex and age. The risk of false negative result was again associated with age (OR of 1.56 for 10 years younger patients, 95% CI: 1.51-1.61), and sex (female to male OR of 2.01, 95% CI: 1.69-2.39; supplementary table 3). Dividing the patients into 2 age groups of similar size (<40 and ≥40, table 1), we found that FNR during this initial period was significantly higher for the younger age group (p-value 0.0002, Fisher’s exact test, OR=1.41, 95% CI: 1.15-1.72; figure 2B). Similarly, the FNR during the later period (days 6-24) also significantly decreased with age (p-value 0.02, Fisher’s exact test).

Based on previous reports of viral load differences between males and females, among age groups and along disease progression^5,16–19,21,29–34^, we hypothesized that differences in FNR across demographic factors and disease progression may stem from changes in viral load, which would be reflected in the measured Ct values. To test this hypothesis, we first tested for associations of Ct values of the three viral genes (N gene, E gene and RdRp) and the internal control gene (IC) with patient age and sex and number of days after diagnosis (figure 3, supplementary figure 1). Indeed, a linear regression model revealed positive correlation of the Ct of viral genes with the number of days after diagnosis, and negative correlation with age and sex (male; Methods: *‘Linear regression’*, supplementary table 4). An opposite association was found with the IC gene, in agreement with within-tube competition for reagents between the multiplexed reactions (supplementary figure 1C)^35^. The viral load association with demographics and time, therefore, mirrored the associations found for the FNR.

**Figure 3.**
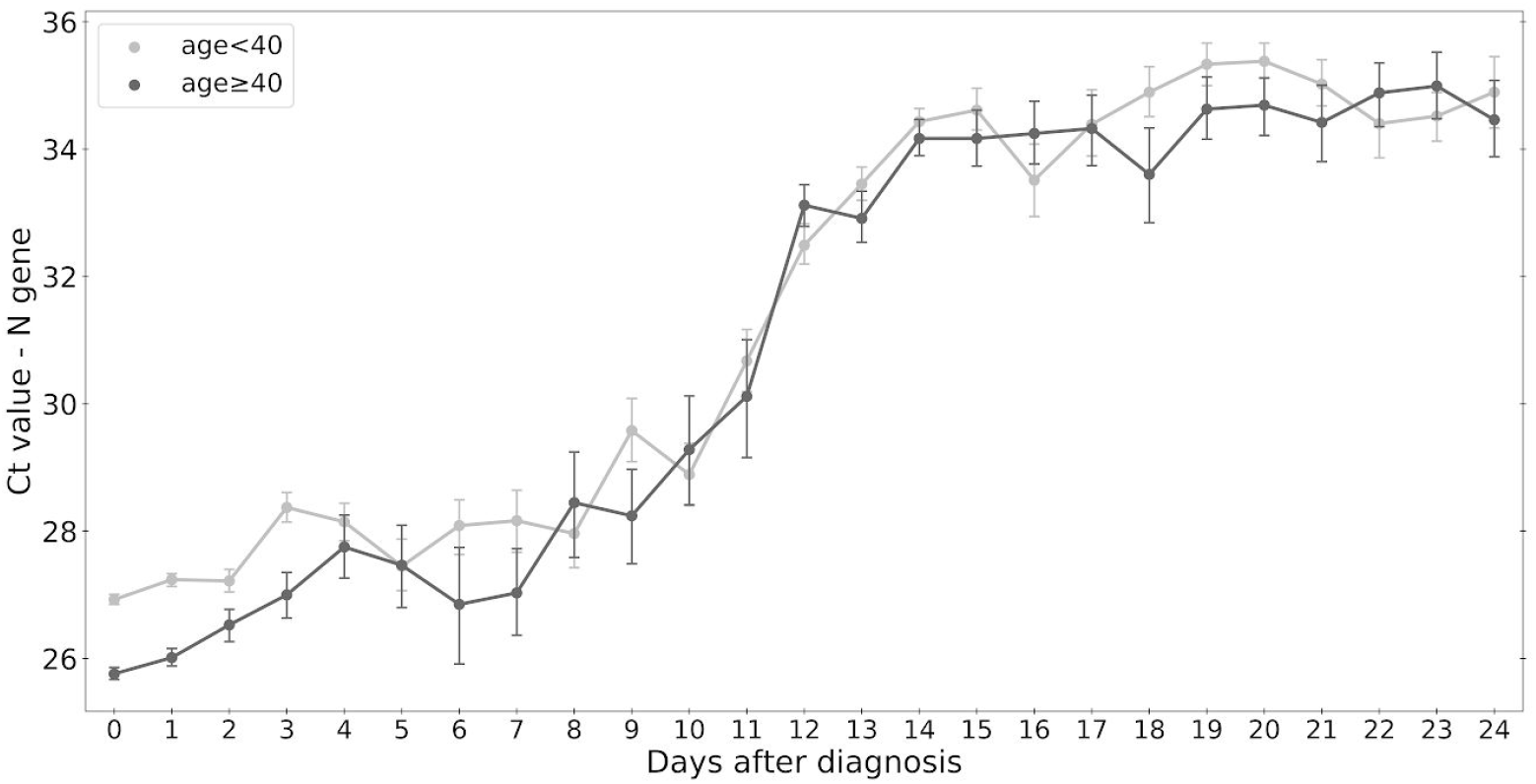
Differential change in Ct value of N gene along time after day of diagnosis for different age groups. In the first 4 days after diagnosis, Ct values of N gene are lower for older patients (age ≥40, light grey) than for younger patients (age <40, dark grey). Error bars indicate SE.

Finally, we tested more directly for association of false-negative rate with viral load at the individual patient level. Since Ct values are not available for false-negative results, we used as a proxy the Ct values of the next positive result. Comparing the distribution of Ct values of positive test results following false-negative tests with, as a control, the Ct values of positive test results following true-positive tests, we found that indeed false-negative results are associated with reduced viral load for all three viral genes (figure 4 and supplementary figure 2; Mann-Whitney U test; p-value of genes, respectively). 4.6 * 10^−4^, 9.0 * 10^−5^, 7.7 * 10^−4^ for N, E and RdRp

**Figure 4.**
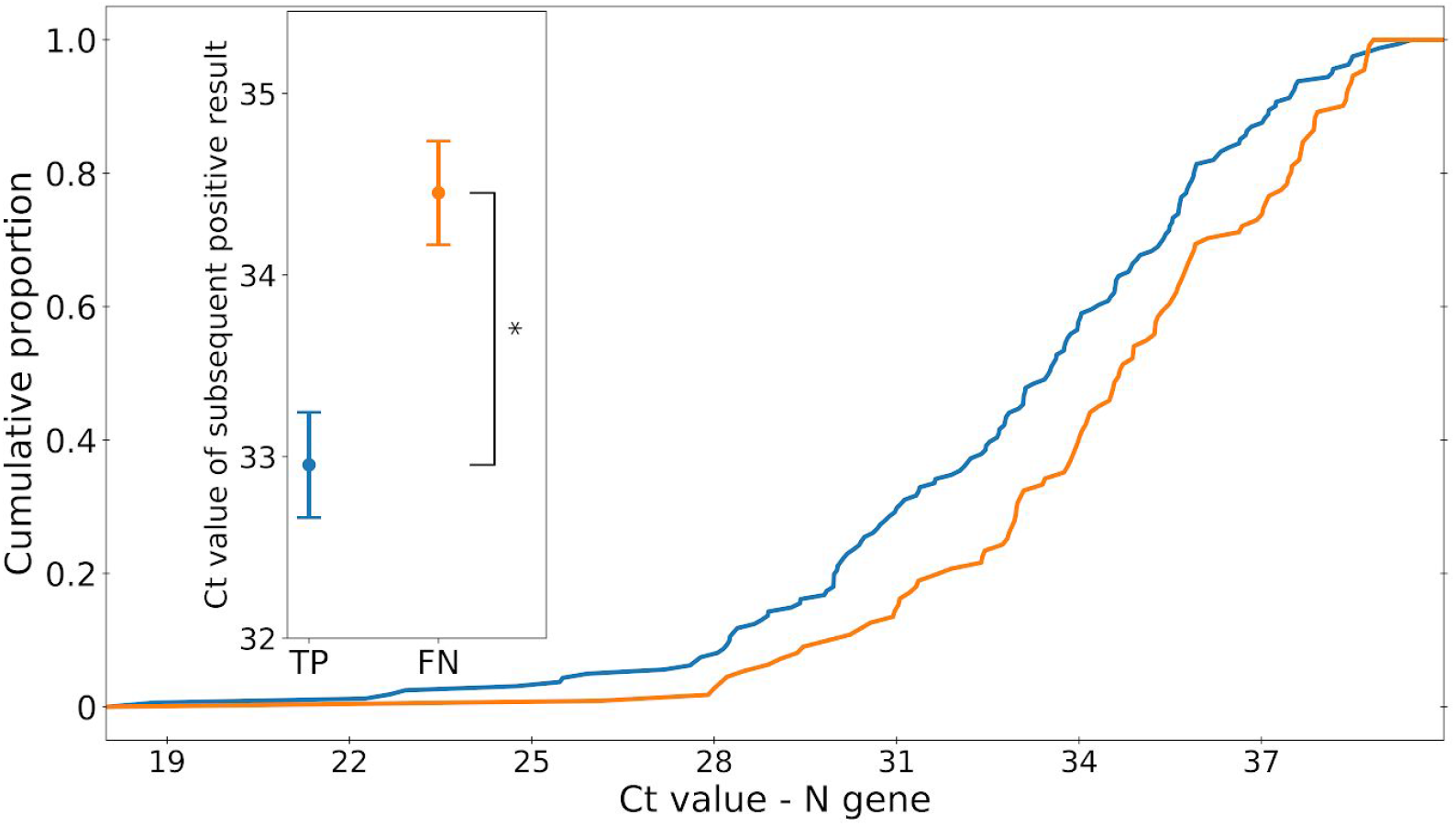
Difference between Ct values of N gene of positive results following FN and TP. Positive test results which followed a false-negative result had a higher Ct value than positive test results following false-negative results. * p-value<0.001. Error bars indicate SE.

## Discussion

Our analysis of large dataset of electronic health records of COVID-19 patients showed that while on average the FNR is about 23%, consistent with past measurements, this rate varies strongly with age, sex and time after diagnosis. At the first few days following diagnosis, the FNR is only 10% on average and even lower for men and older patients. Combining these data with raw fluorescence measurements of RT-qPCR tests for the presence of SARS-CoV-2 genes provides evidence that false-negative rates stem from low viral loads at the single-patient level.

Our study has several limitations. First, we treat all positive tests as true positive. While errors may occur, the rate of false-positive results is very low^2–5^ and we do not expect it to significantly affect our results. Future studies can further improve the reliability of confirmation of positive cases by combining PCR test results with serology tests. Second, we treat negative results at the end of test series as ‘true-negative’, while it is possible that if the test series were continued additional positive tests might have been detected. Again, we do not expect this to significantly affect our results: most series in our study end with two consecutive negative results, and the chances for two consecutive false-negative tests are very low. Moreover, this bias will mostly affect the calculated false-negative rate at later days after diagnosis. Third, as viral loads after infection may first increase and only later decrease, it is possible that false-negative rates follow an opposite pattern: first decreasing and only later increasing. Analysing our cohort, we could only identify the later phase of increasing false-negative rate. However, it is possible that with different cohorts or inclusion criteria, both phases can be observed. Fourth, it will be interesting to see how changing the way Ct is calculated can fine-tune the way positive and negative results are determined based on conflicting results of the genes. Finally, we emphasize that most of the patients in the study cohort were symptomatic; therefore, our results may not represent the false-negative rate for asymptomatic patients.

Despite these limitations, our results provide important epidemiological and clinical input as to the patient specific sensitivity of tests, with important implications for epidemiological policy making, patient-specific quarantine decisions, and disease prevention and control. In particular, they underscore that the risk of false-negative at the very early days following diagnosis might be lower than previously thought, reinforcing the usefulness of these tests for isolation-entry decisions. At the same time, they suggest that tests taken at later time points are less reliable and may therefore be less adequate for isolation-release decisions.

## Data Availability

Data is available upon reasonable request.

---

**Supplementary Table 1.** Series ending results.

| # of consecutive true negative results | # of patients | frequency (%) |
| --- | --- | --- |
| 1 | 2,489 | 32 |
| 2 | 4,237 | 54 |
| 3 or more | 1,146 | 14 |

**Supplementary Table 2.**
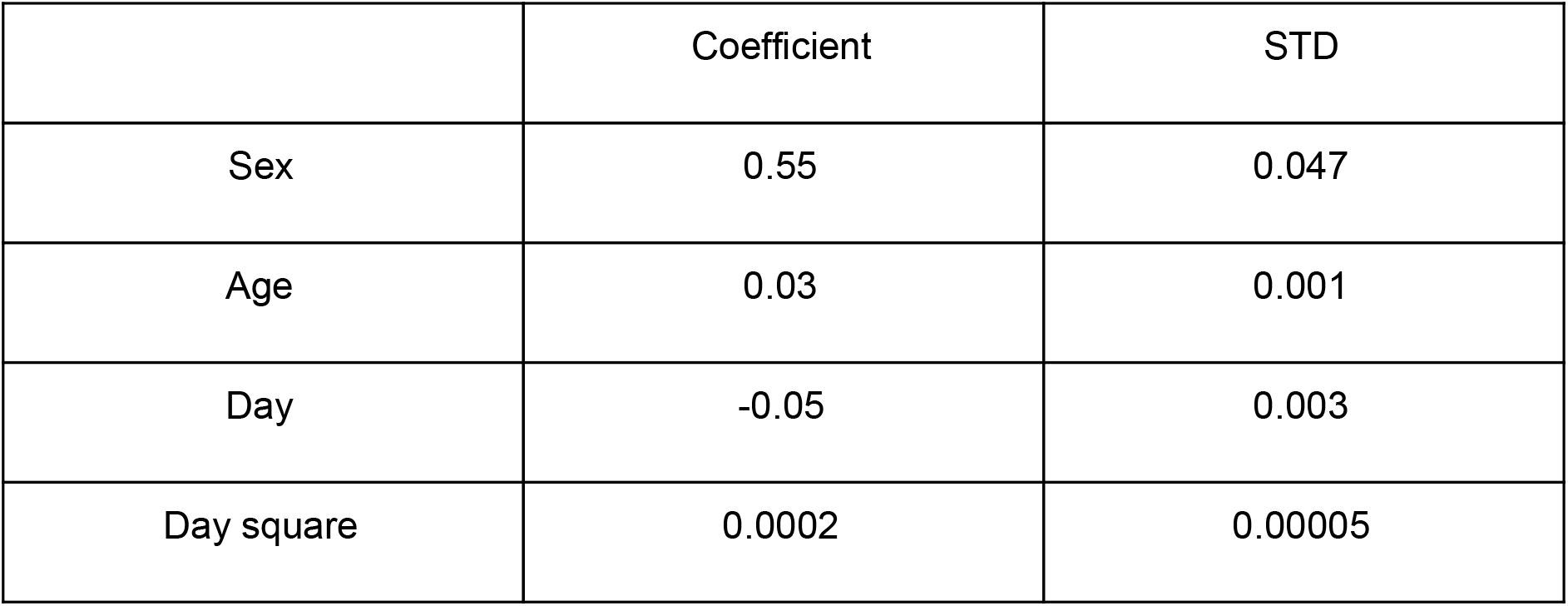
Logistic regression coefficients for a false-negative result.

**Supplementary Table 3.** Logistic regression coefficients for a false-negative result - days 0-5.

|  | Coefficient | STD |
| --- | --- | --- |
| Sex | 0.70 | 0.09 |
| Age | 0.04 | 0.002 |

**Supplementary Table 4*.** Linear regression coefficients for Ct value.

| Gene | N | E | RdRp | IC |
| --- | --- | --- | --- | --- |
| Constant | 27.51<br>[27.34-27.68] | 24.25<br>[24.07-24.43] | 25.90<br>[25.72-26.07] | 27.45<br>[27.32-27.57] |
| Sex coefficient | -0.24<br>[-0.40-(-0.09)] | -0.19<br>[-0.35-(-0.03)] | -0.17<br>[-0.32-(-0.01)] | 0.06<br>[-0.05-0.17] |
| Age coefficient | -0.03<br>[-0.04-(-0.03)] | -0.03<br>[-0.04-(-0.03)] | -0.03<br>[-0.04-(-0.03)] | 0.02<br>[0.01-0.02] |
| Day coefficient | 0.45<br>[0.43-0.46] | 0.50<br>[0.48-0.51] | 0.49<br>[0.47-0.50] | -0.15[-0.16-(-0.14)] |

\*coefficient [95% CI]

**Supplementary Figure 1.**
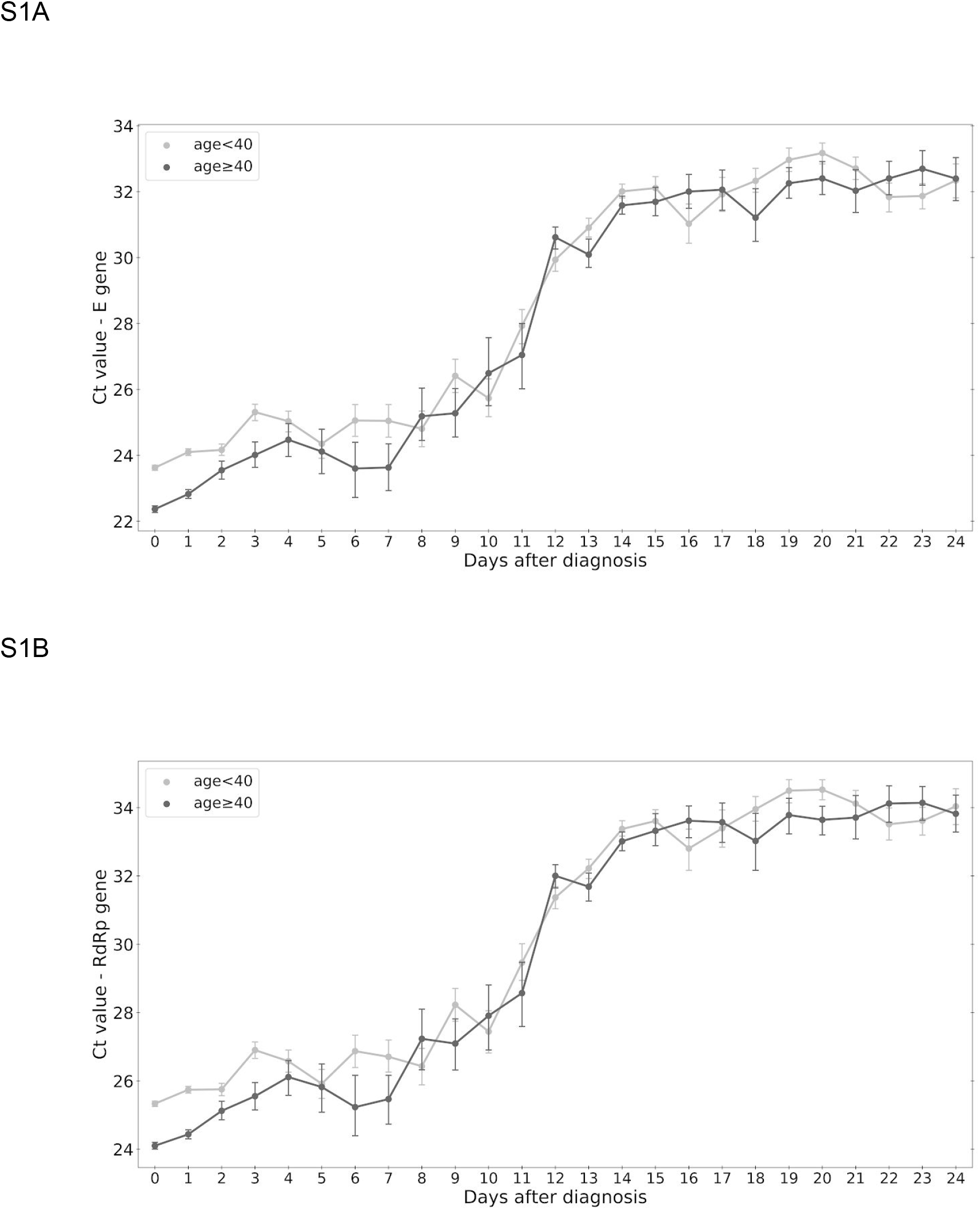
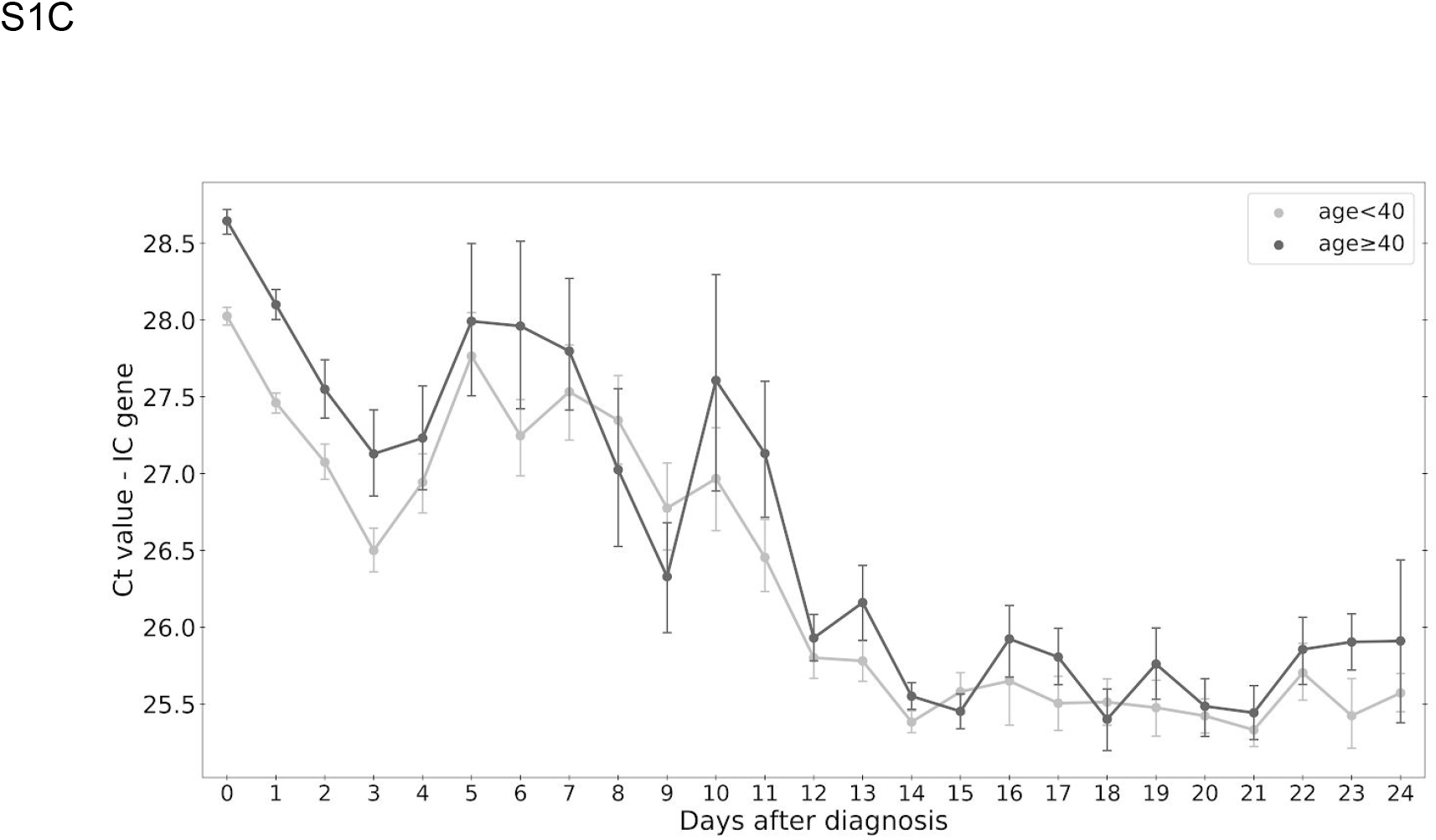
Differential change in Ct value along time after day of diagnosis for different age groups. In the first 4 days after diagnosis, Ct values of E gene (**A**), RdRp gene (**B**) and N gene (main text, Figure 3) are lower for older patients (age ≥40, light grey) than for younger patients (age <40, dark grey). The opposite trend appears for the IC gene (**C**) in agreement with within tube competition for reagents between the multiplexed reactions. Error bars indicate SE.

**Supplementary Figure 2.**
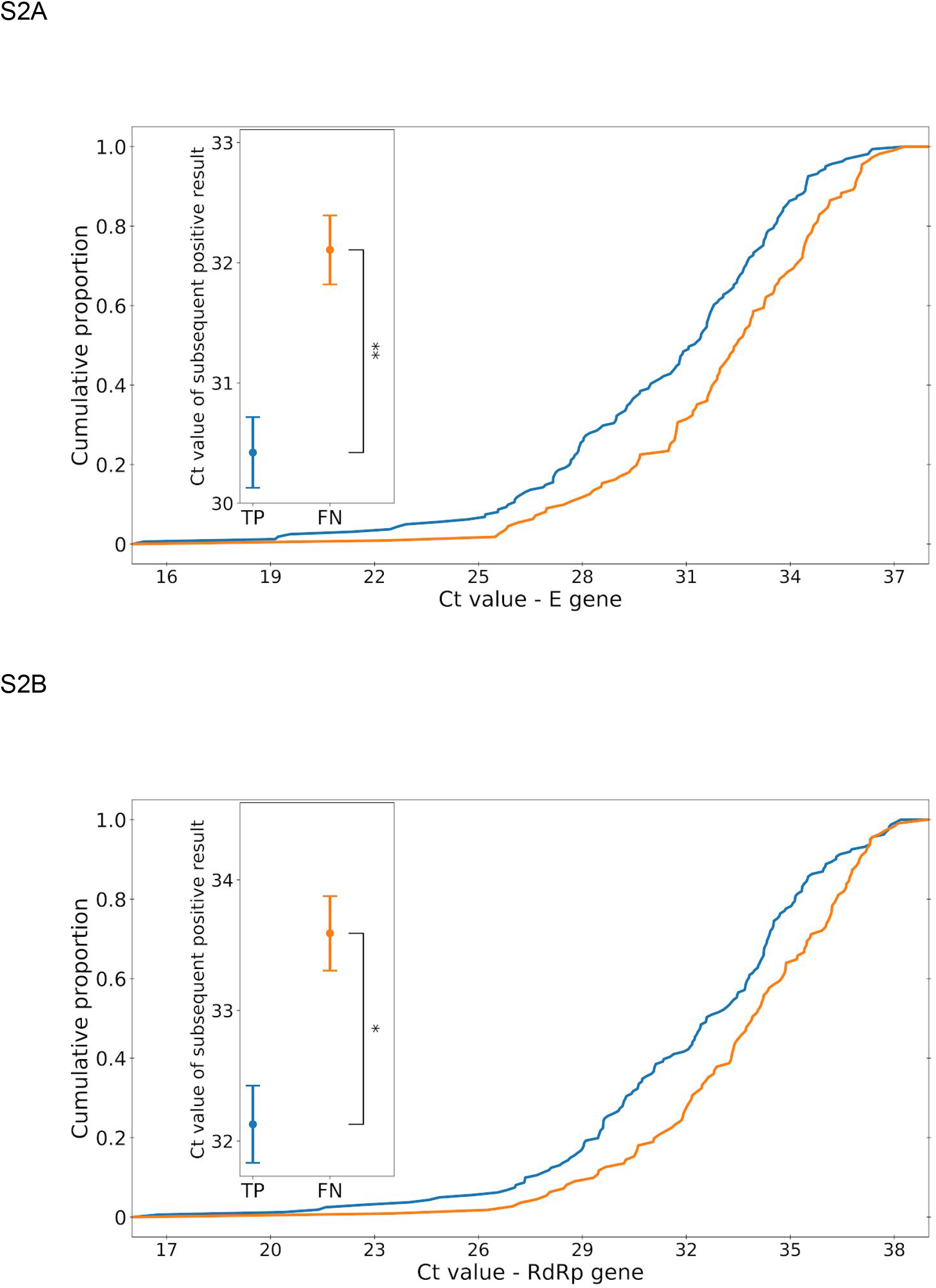
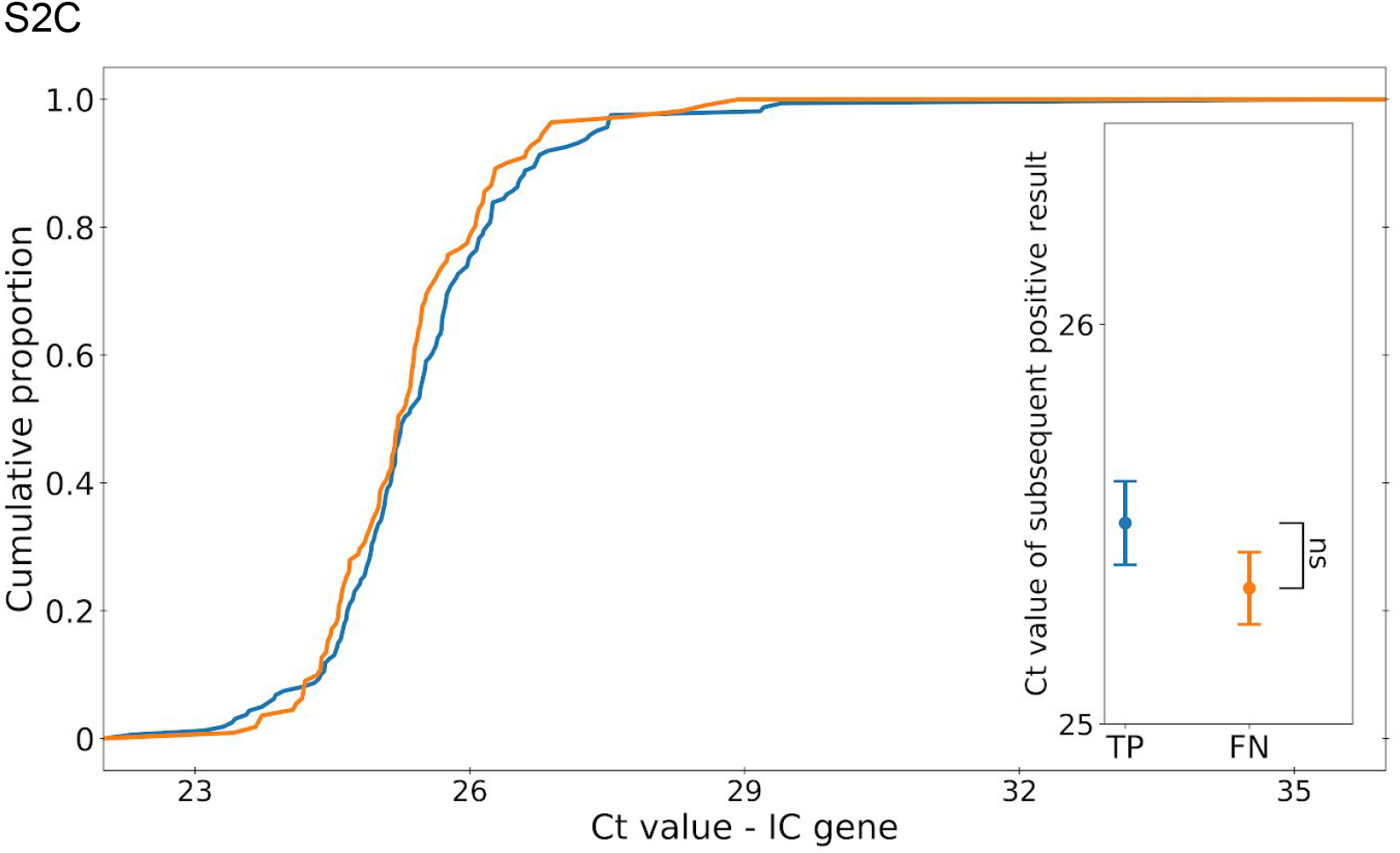
Difference between Ct values of positive results following FN and TP. Positive test results which followed a false-negative result had a higher Ct value, for E gene (**A**), RdRp gene (**B**) and N gene (main text, Figure 4) whereas no significant difference was observed for the IC gene. * p-value<0.001, ** p-value<0.0001. Error bars indicate SE.

